# *Clostridioides difficile* transmission: a compartmental model accounting for environmental spore persistence

**DOI:** 10.1101/2024.04.28.24306515

**Authors:** Jon Edman-Wallér, Kristina Rizzardi, Gunnar Jacobsson, Philip Gerlee

## Abstract

**Objective:** To set up a compartmental model, including environmental spore levels, for *C. difficile* transmission dynamics in a hospital and determine the effect of preventive interventions on infection and colonization rates.

**Design:** Intervention study within a compartmental mathematical modeling framework.

**Setting:** A simulated Swedish 500-bed secondary care hospital.

**Interventions modeled:** Antibiotic stewardship, improved isolation of infected patients, improved general cleaning and disinfection.

**Results:** Antibiotic stewardship had the largest effect on infections, with a 30.6% decrease in infection prevalence. Improved general cleaning and disinfection had the largest effect on colonization (–22.5%) and environmental spore levels (–39.7%). Improved isolation of infected patients had modest effects in comparison.

**Conclusions:** Modeling that includes the dynamics of environmental spores can aid our understanding of *C. difficile* transmission within hospitals. Antibiotic stewardship and improved general cleaning and disinfection showed the largest potential for prevention in our modeled setting.

## Introduction

*Clostridioides difficile* infections (CDI) are among the most common healthcare-associated infections worldwide. Forming spores resistant to ordinarily used disinfectants, the bacteria can spread from infected or asymptomatically colonized patients to the hospital environment and indirectly to new patients. Hospitals harbor both colonized or infected patients at risk of spreading *C. difficile* and patients at risk of acquiring *C. difficile* during their hospital stay. However, in the absence of epidemic strains, only a minority of infections can be traced to transmission from other symptomatic patients. ^1, 2^ Other possible sources of transmission include (but are not limited to) environmental exposition in the community, transmission via healthcare workers (HCW), and transmission from asymptomatic carriers. Interventions to reduce the rate of CDI may be costly and, in some cases, of questionable impact.

*C. difficile* transmission dynamics have been modeled previously, with various versions of adapted compartmental models based on the classic Susceptible, Infected, Recovered (SIR) model, ^3–5^ or agent-based models. ^6–8^ SIR models often assume a rather direct person-to-person transmission, which does not describe the dynamics of *C. difficile* well. Spores from infected or colonized patients are deposited in the hospital environment, where they accumulate if not removed, and can later infect a new patient. Without specific modifications, an SIR model does not model the gradual increase of infections if cleaning and disinfection routines are lacking. When transmission from the environment has been included in previous compartmental models, it has been modeled as a constant transmission source. ^9^ Some agent-based modeling studies have incorporated environmental transmission, ^6, 8^ but include many assumptions of probabilities that are mainly based on expert opinion. In this study, we present a simple, modified compartmental model encompassing a separate compartment for the environmental spore reservoir. We then determine the modeled effects of several possible interventions for reducing hospital CDI incidence, colonization, and environmental spore contamination.

## Methods

### Setting

The hospital in the model was assumed to be a 500-bed secondary-care hospital in Sweden. The model used aggregated compartments with assumptions of homogeneity. Input data from sources similar to our setting were preferred. Sweden has a low epidemic ribotype 027 (RT027) prevalence and a high diversity of strains. ^10^ The number of hospital beds is among the lowest in the OECD. ^11^ Antibiotic consumption is low, ^12^ as is the prevalence and burden of antimicrobial resistance. ^13^ *C. difficile* outbreaks have occurred infrequently, and incidence has declined in recent years. ^10^ Still, CDI is a common healthcare-related infection with an incidence of 61 cases per 100,000 in 2021. ^14^ Routines for infection control of CDI patients vary within the country but generally include care in a single room with a private bathroom until two days after symptom resolution; hand-washing for healthcare workers followed by alcoholic hand rub after contact with the patient or their surroundings; personal protective equipment based on contact precautions; daily disinfection of patient-near surfaces with a disinfectant with or without sporicidal effect; and thorough cleaning of the room at discharge.

### Model

The model was adapted from the work by Yakob et al. ^3^ with the following changes: Firstly, we disregarded the exposed compartment, into which unexposed patients enter upon contact with *C. difficile* spores, and which they leave for the colonized compartment after a predefined period. In Yakob et al., this compartment played an essential role since they investigated the effect of CDI screening upon hospital admission, a procedure that can only detect colonized patients. The interventions we consider do not rely on the exposed/colonized distinction, and we could let patients flow directly from unexposed to colonized. Secondly, we did not consider separate compartments for patients who recently have taken antimicrobials and thus are vulnerable. This was motivated by the fact that while antibiotics increase the risk of CDI, antibiotic treatment is not a prerequisite for infection. Lastly, to more accurately capture the fact that transmission occurs via the spores in the environment, we included an explicit compartment for environmental spores. This environment should be interpreted broadly and includes, for instance, the hands of healthcare workers (HCWs). HCWs were not modeled as separate compartments as they are infrequently infected or colonized. ^15^ We thus ended up with the following compartments in our model: patients unexposed to CDI (U), patients that have become colonized with *C. difficile* spores (C), patients that are infected (I), and those that are in recovery from an infection (R). Additionally, there is an environmental compartment (E) with spores instead of patients.

We consider a hospital with a fixed number of beds (N). The flow of newly admitted patients occurs into the U, C, and I compartments, where the proportion admitted into each compartment is controlled by the parameters ε_U_, ε_C_, and ε_I_. The outflow of patients occurs from the U, C, and R compartments, is equal for the three compartments and given by κ. Spores in the environment are produced by colonized, infected, and recovered patients at rates α_C_, α_I_, and α_R_, respectively, which increase the spore reservoir level E. Cleaning of the hospital environment reduces the number of spores at a rate δ. The number of new colonizations per time unit depends on the presence of unexposed patients and spores in the environment. It is controlled by the parameter 𝛽 and the environmental spore reservoir E. The variable E is given in a non-dimensional normalized form, where the spore level at equilibrium in the baseline scenario equals one (see below). Colonized patients become infected at a rate 𝜃, and infected patients become recovered at a rate 𝜌. No recurrences occur within the hospital, so recovery is the last possible stage for patients before they are discharged.

The following system of coupled ordinary differential equations describes the model:

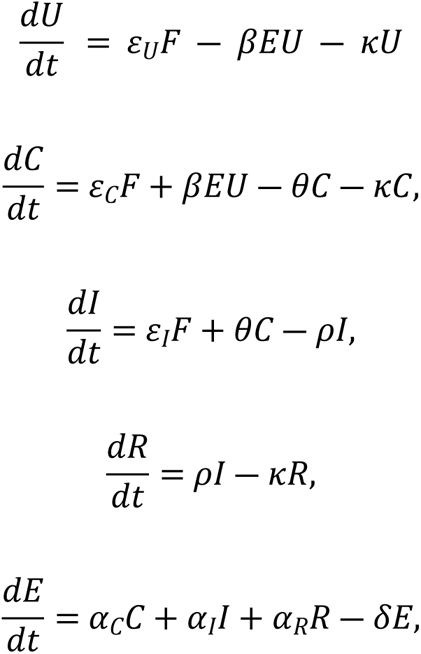

where 𝐹 = 𝜅(𝑁 ― 𝐼) is the outflow of patients, and 𝑁 = 𝑈 + 𝐶 + 𝐼 + 𝑅 is the total number of patients. A schematic description is given in Figure 1, which also shows the flow of patients between the compartments and the flow of spores to and from the environmental compartment E. The model was solved numerically using the Isoda-solver as implemented in the SciPy-method odeint. The initial condition was set to (𝑈,𝐶,𝐼,𝑅,𝐸)(𝑡 = 0) = (0.95𝑁,0.05𝑁,0,0,0) and, to determine the equilibrium state, the model was solved for 1000 days. We tried a range of initial conditions and found that the equilibrium was insensitive to the exact choice of initial conditions. The code is available at https://github.com/philipgerlee/CDITransmission.

**Figure 1.**
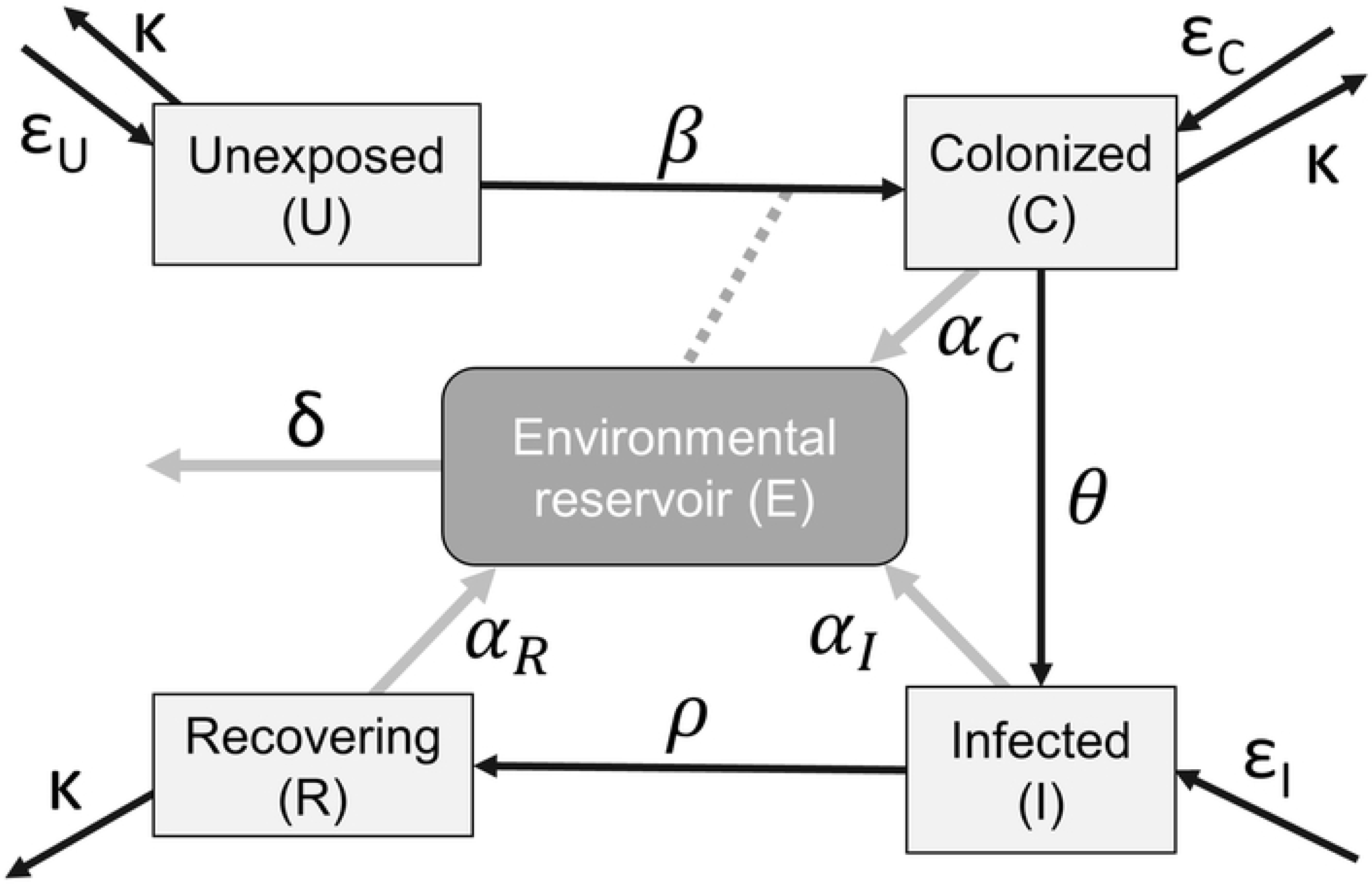
Schematic description of the model. Boxes correspond to compartments and black arrows indicate patient flow between compartments. Spore flows are represented by grey arrows. The dotted line represents an increased patient flow from unexposed to colonized (C) dependent on the level of spores in the environment (E). For interpretation of symbols, see *Table 1*.

**Table 1.** Model variables and parameter values. CDI: *C. difficile* infection.

| *Model parameters* |  |  |  |  |
| --- | --- | --- | --- | --- |
| Parameter | Interpretation | Input value range | Baseline model value | Comments and references |
| N | Number of patients in the hospital |  | 500 | Aimed to represent a secondary-care hospital in Sweden |
| F | Inflow of patients | | $\kappa(N-I)$ | Balanced against outflow for a constant N |
| $\epsilon_U$ | Proportion unexposed at admission | | 0.949 | $1 - (\epsilon_C + \epsilon_I + \epsilon_R)$ |
| $\epsilon_C$ | Proportion colonized by toxigenic *C. difficile* at admission | 0.019-0.115 | 0.05 | 1.9%,<sup>18</sup> 5.2%,<sup>19</sup> 11.5%<sup>20</sup> |
| $\epsilon_I$ | CDI prevalence at admission | 0.0008-0.002 | 0.001 | Incidence of CDI in western Sweden in 2016: 0.8/1000 days of care<sup>10</sup> implies $(0.8/\kappa)/1000$ admissions = 0.004, of which a minority (range 20–50%) occur before hospital admission.<sup>21</sup> |
| $\kappa$ | Rate of discharge from hospital for U, C, R | 0.17-0.26 | 0.2 | Mean length of stay at Swedish hospitals = 3.9 days, <sup>22</sup> 5.7 days. <sup>23</sup> |
| $\beta$ | Rate unexposed -> colonized | 0.016-0.021 | 0.017 | Net colonized during their stay: 8.5%, <sup>24</sup> 6.4%<br><sup>20</sup><br>Divided by $1/\kappa$ to obtain rate per day: 2.1%, <sup>24</sup> 1.6% <sup>20</sup> |
| $\theta$ | Rate colonized -> infected | 0.003-0.006 | 0.004 | Incidence of CDI in western Sweden in 2016: 0.8/1000 days of care, <sup>10</sup> of which a majority (range 50–90%, 0.0004–0.007) occurred among hospitalized patients. <sup>21</sup><br>As all infections occur among colonized patients, this rate is divided by %C (at equilibrium = 0.121) for a range of 0.003-0.006. |
| $\rho$ | Rate infected -> recovered | 0.14-0.33 | 0.16 | 1-3 days from symptoms to treatment, 2-4 days<br>from treatment to symptom resolution<sup>25, 26</sup><br>for vancomycin, 4.6 days for Metronidazole. <sup>27</sup><br>Metronidazole is still widely used in Sweden, with a rate of treatment failure around 12% <sup>28</sup>, after which vancomycin is usually prescribed and clinical cure is delayed. This makes the lower end of the range most plausible in our setting. |
| $\delta$ | Clearance rate of spores from the environment | 0.01-0.03 | 0.0175 | Transmission between patients via the hospital environment has been traced several months from donor to recipient. <sup>1</sup><br>Persistence of spores in the hospital environment up to five months. <sup>29</sup> |
| $\alpha_I$ | Relative increase of E per day caused by an | - | 0.001 | Value determined by feedback of model output (see Methods). |

|  | individual infected patient |  |  |  |
| --- | --- | --- | --- | --- |
| $\alpha_C$ | Relative increase of E per day caused by an individual colonized patient | 0.08-0.65 * $\alpha_I$ | 0.00025 | 0.25 * $\alpha_I$<br>Relative frequency of excess hospital contamination (rates above uncolonized controls) in colonized versus infected patients range from 0.08-0.65 depending on room area.<br>30 |
| $\alpha_R$ | Relative increase of E per day caused by an individual patient in recovery | 0.02-0.2 * $\alpha_I$ | 0.0001 | 0.1 * $\alpha_I$<br>Low frequency of *C. difficile* in stools during treatment. 31 |
| *Model state variables for internal and external consistency* |  |  |  |  |
| Variable | Interpretation | Target value | Baseline model output value | Comments and references |
| E | Hospital spore reservoir at equilibrium | 0.99-1.01 | 0.998 |  |
| U | Proportion of unexposed patients | - | 435 patients (87.1%) |  |
| C | Proportion of colonized patients (prevalence of colonization) at equilibrium | 5-13.5% | 61 patients (12.1%) | $> \varepsilon_C$ (0.05) – more than the proportion colonized at admission<br>$< \varepsilon_C + \beta/\kappa$ (0.135) – less than the proportion colonized at discharge |
| I | Proportion of infected patients (prevalence of CDI) at equilibrium | 0.2-0.5% | 2 patients (0.43%) | 0.36% (own hospital data)<br>0.2%, <sup>16</sup> 0.5% <sup>17</sup> |
| R | Proportion of recovering patients | - | 2 patients (0.34%) |  |

### Determination of parameter values

First, a literature search was performed for each model parameter to find evidence for a range of plausible values. The values of α_C_, α_I,_ and α_R_, representing the contribution of spores from individual patients per day to the total environmental reservoir of spores, were difficult to estimate based on the literature and were only estimated relative to each other at this stage. Second, after setting arbitrary values for the α parameters and mid-range values for all other parameters, the model was solved numerically, and the proportion of colonized and infected patients in equilibrium was determined. Third, these values were compared to expected colonization levels (5-13.5%, see Table 1) and CDI prevalence (0.2-0.5%, ^16, 17^, own data) among hospitalized patients in our setting. When the values were not within these ranges, the parameters were changed manually within their respective range until the model resulted in colonization and CDI prevalence within the expected ranges. The α parameters were adjusted first, and then the other parameters. Fourth, for the model to be internally consistent at baseline, βE (in the model) should be equal to our estimation of β at baseline, i.e., E should be = 1. This is because β is estimated based on studies close to our setting, with a certain level of environmental spores. Thus, the estimation of β is actually an estimation of βE. Parameter values, especially the α values (contribution to environmental spores by patients) and the δ value (clearance of spores from the environment), were adjusted within their ranges until the model resulted in E = 1 at equilibrium as well as expected levels of colonization and infection. Parameters, parameter ranges, final baseline model parameters, and outputs in the equilibrium state are presented in Table 1.

### Interventions

Having built a model representing our current setting, we simulated three possible CDI-reducing interventions to estimate their expected effect on the environmental spore burden and the prevalence of *C. difficile* infection and colonization. The effects of the interventions were studied on the scale from no change to very ambitious while still within the possible.

### Intervention 1: Antibiotic stewardship program

A recently updated meta-analysis ^32^ of the increased risk for CDI in hospitalized patients for different classes of antibiotics was used to estimate the odds ratios (ORs) for different antibiotic classes. The point estimates of ORs in the study were used except when confidence intervals overlapped 1; in that case, the OR was set to 1. Swedish data on hospital antibiotic use in 2021 ^33^ in defined daily doses (DDD) for each category was multiplied by 𝑂𝑅 ― 1 to estimate the increase of CDI cases attributable to the antibiotic in the baseline model (table 2). Estimates of the increased CDI risk attributable to different antibiotic classes combined with Swedish data on hospital consumption of antibiotics (Table 2) resulted in an estimation that antibiotics with an OR >1 increase the number of cases by 44.1%, implying that 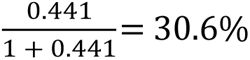 of cases could be prevented if low-risk antibiotics or no antibiotics replaced high-risk antibiotics. To model this intervention, we varied the θ parameter from 0.004 to as low as 0.00233 to achieve an up to 30.6% decrease in the infected population I at equilibrium.

**Table 2.** Antibiotics: consumption and increased *C. difficile* infection (CDI) risk. DDD: Defined daily dose. OR: Odds Ratio.

| Antibiotic class | DDD/100 patient-days | Share of total DDD/100 patient-days | OR | OR-1 | Increase of CDI cases attributable to the antibiotic class (DDD share * (OR – 1)) |
| --- | --- | --- | --- | --- | --- |
| Cephalosporins | 8.7 | 11.5% | 1.8 | 0.8 | 9.0% |
| Beta-lactamase<br>resistant<br>penicillins | 17.1 | 22.5% | 1.4 | 0.4 | 7.4% |
| Carbapenems | 3.6 | 4.7% | 2,6 | 1.6 | 7.4% |
| Penicillins with<br>extended<br>spectrum | 6.1 | 8.0% | 1.8 | 0.8 | 6.2% |
| Combinations of<br>penicillin | 10.6 | 14.0% | 1.4 | 0.4 | 6.0% |
| Fluoroquinolones | 6.8 | 9.0% | 1.34 | 0.34 | 3.0% |
| Glycopeptides | 1.4 | 1.8% | 1.9 | 0.9 | 1.7% |
| Lincosamides | 1.9 | 2.5% | 1.56 | 0.56 | 1.4% |
| Macrolides | 1.2 | 1.6% | 1.2 | 0.2 | 0.3% |
| Beta-lactamase<br>sensitive<br>penicillins | 7.1 | 9.4% | 1 | 0 | 0% |
| Tetracyclins | 4.1 | 5.4% | 1 | 0 | 0% |
| Trimethoprim-<br>sulfamethoxazole | 3.6 | 4.7% | 1 | 0 | 0% |
| Aminoglycosides | 2.8 | 1.1% | 1 | 0 | 0% |
| Other | 2.9 | 3.8% | 1 | 0 | 0% |
| Total | 75.9 | 100.0% |  |  | 44.1% |

**Table 3.** Maximal effects of interventions. Grey background indicates deviation from the baseline value. For definitions of parameters, see Table 1.

| Parameter | Baseline value | Antibiotic stewardship | Increased CDI isolation | Increased cleaning and disinfection | All interventions combined |
| --- | --- | --- | --- | --- | --- |
| N | 500 | 500 | 500 | 500 | 500 |
| F | $\kappa(N-I)$ | $\kappa(N-I)$ | $\kappa(N-I)$ | $\kappa(N-I)$ | $\kappa(N-I)$ |
| $\varepsilon_U$ | 0.949 | 0.949 | 0.949 | 0.949 | 0.949 |
| $\varepsilon_C$ | 0.05 | 0.05 | 0.05 | 0.05 | 0.05 |
| $\varepsilon_I$ | 0.001 | 0.001 | 0.001 | 0.001 | 0.001 |
| $\kappa$ | 0.2 | 0.2 | 0.2 | 0.2 | 0.2 |
| $\beta$ | 0.017 | 0.017 | 0.017 | 0.017 | 0.017 |
| $\theta$ | 0.004 | 0.00233 | 0.004 | 0.004 | 0.00233 |
| $\rho$ | 0.16 | 0.16 | 0.16 | 0.16 | 0.16 |
| $\delta$ | 0.0175 | 0.0175 | 0.0175 | 0.02275 | 0.02275 |
| $\alpha_I$ | 0.001 | 0.001 | 0.0001 | 0.001 | 0.0001 |
| $\alpha_C$ | 0.00025 | 0.00025 | 0.00025 | 0.00025 | 0.00025 |
| $\alpha_R$ | 0.0001 | 0.0001 | 0.0001 | 0.0001 | 0.0001 |
| *Output* |  |  |  |  |  |
| Environmental spores | 0.998 | 0.936 | 0.781 | 0.601 | 0.484 |
| Unexposed | 87.1% | 87.6% | 88.6% | 90.0% | 90.9% |
| Colonized | 12.1% | 11.8% | 10.7% | 9.39% | 8.63% |
| Infected | 0.43% | 0.30% | 0.39% | 0.36% | 0.25% |
| Recovering | 0.34% | 0.23% | 0.31% | 0.29% | 0.20% |

| *Output, change relative to baseline* |  |  |  |  |  |
| --- | --- | --- | --- | --- | --- |
| Environmental spores | - | -6.2% | -21.8% | -39.7% | -51.5% |
| Unexposed | - | +0.6% | +1.8% | +3.3% | +4.4% |
| Colonized | - | -2.5% | -12.2% | -22.5% | -28.9% |
| Infected | - | -30.6% | -8.6% | -16.0% | -41.5% |
| Recovering | - | -30.6% | -8.6% | -16.0% | -41.5% |

### Intervention 2: Enhanced efforts on reducing the spread from infected patients to the environment

For this intervention, we imagined that the transmission of spores from infected patients would be virtually eliminated. For instance, infected patients could be promptly moved to a separate building not part of the main hospital after diagnosis. We modeled this as a reduction of α_I_ up to 90%, from 0.001 to 0.0001, with all other variables being constant.

### Intervention 3: General increased removal of spores

In the baseline model, spores are removed at a rate δ = 0.0175. This represents the total effect of cleaning, disinfection, hand hygiene, etc., that remove spores from the hospital environment and thus prevents further transmission. While our model cannot determine the individual contribution of each of these practices for spore removal, we can simulate an intervention aimed at various hygienic measures, where the removal of spores increases by a certain amount. We modeled this by increasing δ by up to 30%, from 0.0175 to 0.02275.

### Intervention 4: Bundled intervention

To study the effect of an ambitious bundled intervention with maximal effect aimed at antibiotics and isolation of infected patients as well as general cleaning and disinfection, we combined the three interventions above at their maximal effect.

## Results

### Intervention 1: Antibiotic stewardship program

This intervention resulted in a maximal decrease of colonized patients from 12.1% to 11.8% (–2.5%), a maximal decrease in disease prevalence from 0.43% to 0.30% (–30.6%), and a maximal reduction of environmental spores from 0.998 to 0.936 (–6.2%) (Figure 2A).

**Figure 2.**
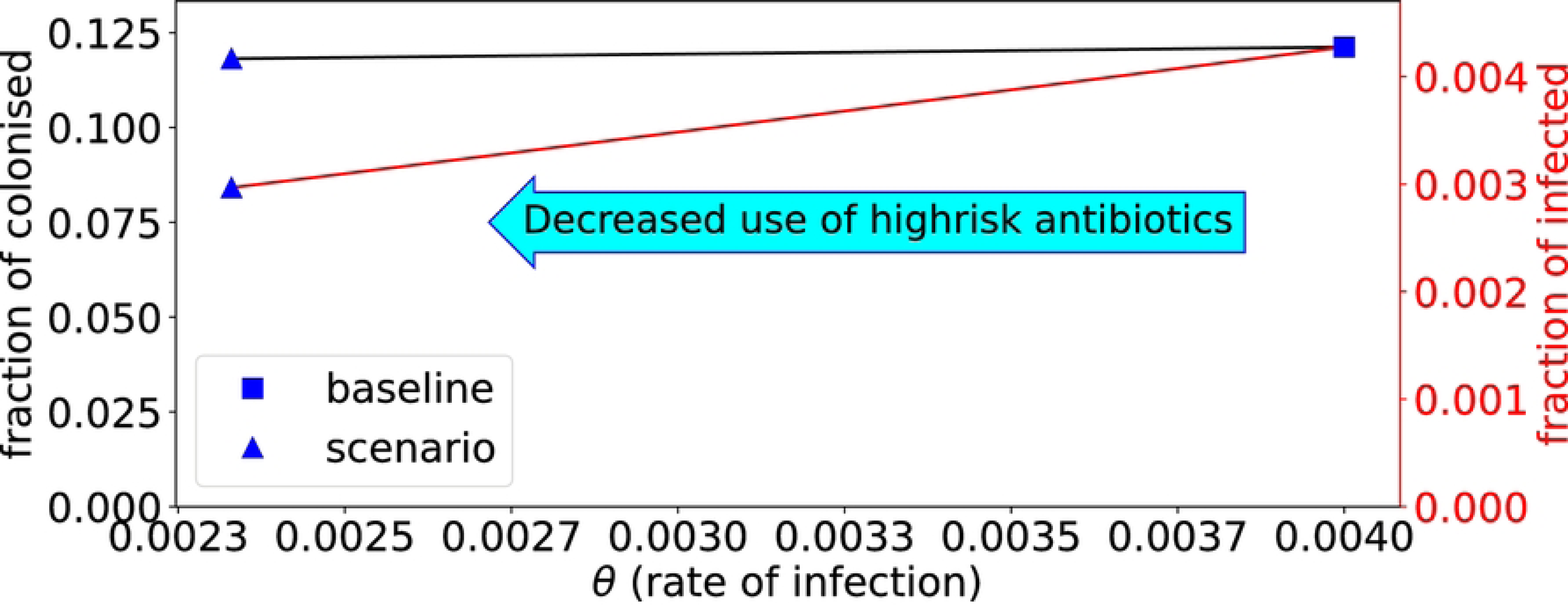
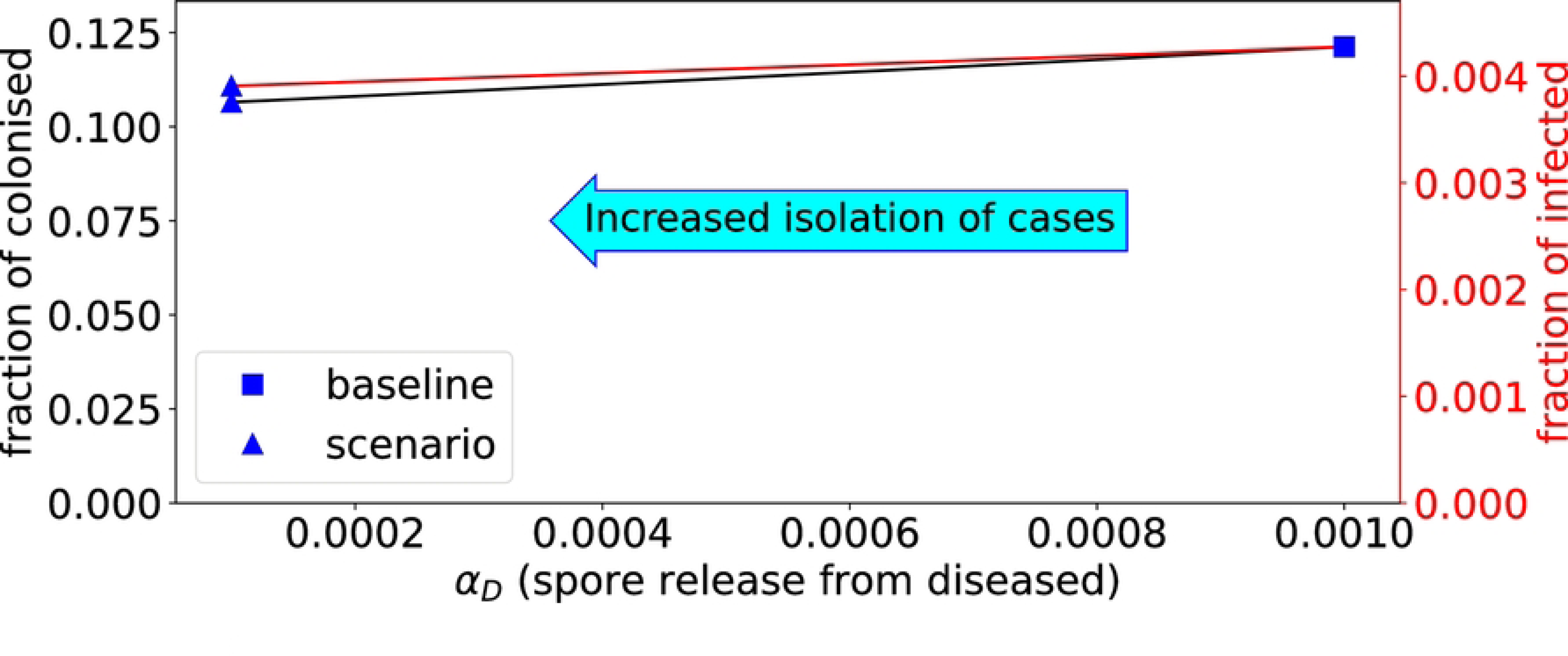
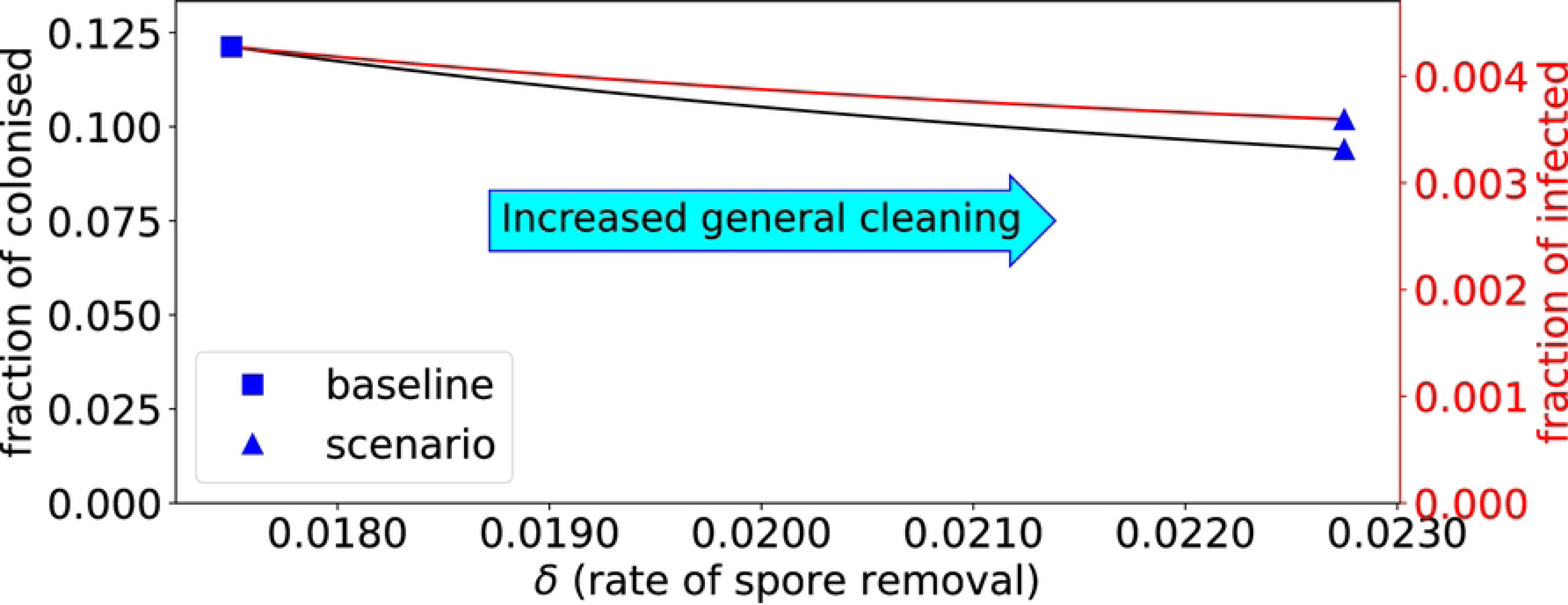
The outcome of the three interventions on the fraction of colonized and infected. A: The effect of decreased use of high-risk antibiotics; B: The effect of case isolation; C: The effect of increased general cleaning and disinfection.

### Intervention 2: Enhanced efforts on reducing the spread from infected patients to the environment

This intervention resulted in a maximal decrease of colonized patients from 12.1% to 10.7% (–12.2%), a maximal decrease of disease prevalence from 0.43% to 0.39% (–8.6%), and a maximal reduction of environmental spores from 0.998 to 0.781 (–21.8%) (Figure 2B).

### Intervention 3: General increased removal of spores

This intervention resulted in a maximal decrease of colonized patients from 12.1% to 9.4% (– 22.5%), a maximal decrease in disease prevalence from 0.43% to 0.36% (–16.0%), and a maximal reduction of environmental spores from 0.998 to 0.601 (–39.7%) (Figure 2C).

### Intervention 4: Bundled intervention

A combination of all three interventions above at their maximum effect resulted in a decrease in colonized patients from 12.1% to 8.6% (–28.9%), a decrease in disease prevalence from 0.43% to 0.25% (–41.5%), and a reduction of environmental spores from 0.998 to 0.484 (– 51.5%) (not included in Figure).

## Discussion

In this study, we aimed to model the transmission dynamics of *C. difficile* infections in a Swedish hospital setting. We modeled three intervention strategies and found that the antibiotic stewardship program had the largest impact on the prevalence of CDI. The impact on environmental spore levels and the prevalence of colonized patients was, however, modest. Vigorous isolation of infected patients moderately decreased colonization rates and environmental spore levels, while the effect on CDI prevalence was modest. Increased reduction of spores by overall cleaning and disinfection greatly reduced colonization rates and environmental spore levels, and moderately decreased CDI prevalence.

The maximum effect of an antibiotic stewardship program, a decrease in cases by 30.6%, might seem on the low side in relation to epidemiological studies on the effects of such programs, which often have achieved CDI incidence decreases of around 50%. ^34^ However, Sweden has a favorable antimicrobial resistance situation, and antimicrobial stewardship programs are already active on national, regional, and local levels. For instance, beta-lactamase-sensitive penicillins with low CDI risk make up almost 10% of the total, almost as much as cephalosporins. ^33^ Thus, it should be expected that the maximal impact of further stewardship measures in our setting is limited. Also, an antibiotic stewardship program might affect the progression from unexposed to colonized or lead to changes in lengths of stay, which were not modeled in this study. However, different antibiotics may increase or decrease the risk of colonization, ^20^ and antibiotics overall did not increase the risk of *C. difficile* colonization in a recent meta-analysis. ^35^

The maximum effect of general cleaning and disinfection was considerable in our study. This contrasts with a recent review and meta-analysis of the effect of environmental cleaning bundles ^36^, which concluded that such interventions seem to have little or no effect on CDI incidence. However, the bundles in the included studies may have been ineffective in increasing the overall reduction rate of environmental spores. Thus, our model implies that if we could substantially increase the reduction rate of spores, this would significantly affect CDI incidence. However, still needs to be determined what interventions would be needed to increase this rate substantially.

Focusing on isolating patients had modest effects in our model, especially on disease prevalence. In this scenario, colonized patients continue contributing spores to the environmental reservoir. Asymptomatic patients as important sources of *C. difficile* infections have been discussed in the literature in recent years. ^37–39^ While testing for colonization and isolating these patients may be practically unfeasible and ethically questionable, our results suggest that more effective cleaning and disinfection practices for all patients may be a way to reduce the risk of transmission from colonized patients; this, however, depends on these practices to increase the riddance of spores to a meaningful degree.

A main strength of our study is that the dynamic effects of the environmental spore reservoir are modeled in a way that, as far as we know, has not been performed previously. We have also calibrated the inputs to ensure that the model produces output that is plausible for our chosen setting and, as far as possible, based our assumptions on scientific data rather than expert opinion. While this study was performed with a Swedish hospital in mind, the model could easily be adjusted to mirror other settings.

Our study has limitations. No model is better than its assumptions, and for some parameters (e.g., the relations between the different α parameters), it was difficult to find clear scientific support for a given level. The model is also a highly simplified version of a hospital. It does not consider movements of patients within the hospital building, division into different wards, or movement of HCWs. The interventions were modeled in a simplified way as well. These shortcomings could be addressed by considering a more complex, potentially agent-based model, but such an approach requires detailed knowledge about model parameters.

We conclude that, according to our model, antibiotic stewardship measures have the greatest potential to decrease CDI incidence in our setting. Cleaning and disinfection efforts, implemented in a way that substantially decreases the environmental spore reservoir, have the greatest potential to prevent in-hospital acquisition of *C. difficile*.

## Data Availability

https://github.com/philipgerlee/CDITransmission

## Acknowledgements

*Financial support*. None reported.

*Potential conflicts of interest*. All authors report no conflicts of interest relevant to this article.

## Notes

### Competing Interest Statement

The authors have declared no competing interest.

### Funding Statement

The author(s) received no specific funding for this work.

